# Enhancing PET/CT Assessment with Porous 3D Printed Grids– A Pilot Study

**DOI:** 10.1101/2025.04.01.25325022

**Authors:** Sai Kiran Kumar Nalla, Quentin Maronnier, Tala Palchan-Hazan, John A. Kennedy, Olivier Caselles

## Abstract

**Objective:** Phantom experiments are widely used for standardisation in positron emission tomography (PET), but current practices to do not necessarily reflect clinical reality and require meticulous phantom preparation for repeatability. 3D printing can reduce these limitations by optimizing preparatory methods and improving phantom features. This work proposes employing 3D-printed porous grids as an alternative mechanism to emulate targets with contrast.

**Approach:** Acrylonitrile butadiene styrene (ABS) cubic grids (4 cm/side) with varying design characteristics and targets were printed. Grids were immersed in a [^18^F]FDG solution with soap within a conventional phantom. Five consecutive acquisitions were repeated on five different days (Day 0, 1,4-6) using a Discovery MI PET/CT. Target representation index(TRI) and dilution coefficient (DC) were the metrics used for analysis. Friedman test was utilized to study global statistical significance across days.

**Main Results:** PET images resulted in clear demarcation of various contrast regions produced by the dilution grid. Quantitative metrics showed consistent results across trials, confirming robustness. Dilutions achieved (mean±std. dev.) were 1.93±0.14, 2.44±0.2, and 3.01±0.08 vs. 2, 2.5 and 3.33 (theoretical), respectively. Observed TRI were in range of 0.4 – 1.2. Correlation across days was strong (p≥0.67), and days 1 and 5 had the best pairwise comparable results.

**Significance:** 3D-printed grids offer a reliable, reproducible alternative for PET/CT assessment. Dozens of targets with background were produced with a single tracer administration. CT attenuation throughout the phantom mimicked water, giving good PET representation of wall-free targets.

## 1. Introduction

Physical and digital phantoms are typically used for assessment of lesion detection and quantitation across image processing algorithms and imaging systems in nuclear medicine (NM) **[1–3]**. Generally, positron emission tomography / computed tomography (PET/CT) phantoms for target assessment consists of multiple compartments and inserts, such as hollow acrylic spheres, to simulate lesions and surrounding tissue **[4,5]**. By meticulously filling these compartments and inserts with various radiotracer activity concentrations contrasts between different regions can be created. An example of such phantom would be the National Electrical Manufacturers Association (NEMA) PET body phantom which has multiple compartments and six fillable spheres **[6,7]**. While useful for standardization and baseline assessment, these phantom systems fail to replicate the complex anatomical variability and heterogeneity seen in clinical reality **[8]**. Moreover, the number of features/compartments and their mounting positions are fixed for traditional phantom designs thus limiting their customizability. Additionally, existing commercial phantoms such as the Jaszczak, Hoffman brain, and various body or torso phantoms, and their respective attachments provide a wide range of platforms but may cost upwards of several thousand dollars **[9]**. Due to its versatile potential, three-dimensional (3D) printing has been previously reported to address some of these limitations of phantoms used in medical imaging **[10–12]**. In the context of PET/CT applications, 3D printing has been used to create complex anatomical phantoms closer to clinic reality with customizable targets emulating lesions, thus show-casing the improved abilities of modern scanning technology for lesion detectability **[8,9,13–20]**. The two major cost-effective methods of 3D printing, namely fused deposition modelling (FDM) and stereolithography (SLA), have been widely utilized in research for the fabrication of phantoms. Studies using polyjet printing have also been reported, but they come at considerably higher cost **[8]**. From the perspective of target creation (for lesion emulation), spherical inserts similar to acrylic spheres were 3D-printed **[21]**. A few studies also provided some advancements in creating target inserts emulating heterogeneous lesions **[19,20]**. By producing several small compartments within the main fillable target, more intricate radiotracer uptake patterns can be represented. While this is a significant improvement over simpler designs in representing clinical complexity, filling of these multiple small compartments is a challenge in practice. In a few other studies, radioactive targets were created via 3D printing by directly mixing radiotracer into the resin and embedding printed inserts within phantoms **[13,16,17,22,23]**. Though the findings are promising, this requires a shielded environment with specific ambient conditions for printing. Another limitation is its suitability only for long half-life radiotracers, or comparatively high activity concentrations for short half-life radiotracers accounting for the time required for printing and data acquisition. Specifically, for commonly employed short-lived radiotracers in PET imaging, such as Fluorine-18 or Gallium-68, the printed inserts are usable only within a single experimental iteration before becoming unusable due to radioactive decay.

A more practical approach involves using 3D-printed fillable grids that better mimic specific tissue distributions and organ structures analogous to certain practices in radiology **[9,24,25]**. They simplify the ability to model complex activity distributions with a single radiotracer injection, as the internal grid structure modulates radiotracer concentration across various regions, eliminating the need to fill multiple compartments. This minimizes the potential for errors arising from the preparation of solutions with different radiotracer concentrations to obtain varying contrasts **[9]**. Additionally, the design parameters can be adjusted to the spatial resolution of the scanner, the need for cold walls between the regions of interest can be avoided **[24]**. Given a grid spacing of less than the spatial resolution of the PET/CT scanner, such volumes are scanned as if they are homogenous, as the radiotracer infuses the grid pattern. Such SLA grids of multiple contrasts were studied for single photon emission computed tomography / computed tomography (SPECT) **[9,24]**. Monte Carlo simulations were also used to validate that such techniques are compatible with existing practices **[9]**.

While SLA printing offers high precision and a good quality surface, primarily because of the resin properties, the densities are typically higher than that of water making them radiologically less ideal for emulating the body’s attenuation of high energy photons **[9,23]**. FDM uses materials such as thermoplastics, like polylactic acid (PLA) or acrylonitrile butadiene styrene (ABS), which have properties making them more radiologically compatible for hybrid imaging applications **[12,26,27]**. Currently, mainly SLA has been explored for grid-based approaches whereas knowledge of FDM’s performance in such applications is limited. Also, these grid-based approaches have focused on SPECT with little emphasis on PET applications. Additionally, none of these grid studies were based on lesion simulation or target detectability but rather to replicate different regions of specific organ. Though previous studies have confirmed that the CT-based PET attenuation correction (CTAC) works for such 3D printing studies, water neutral attenuation properties which can be achieved by FDM filaments are more ideal as mentioned by **[9,23]**. In this work, we aim to introduce FDM based ABS grids which have are closer to desired neutral properties. The primary objective is to evaluate the ability of the grid designs in creating regions of diverse radiotracer concentration using a single radiotracer injection, while concurrently assessing the consistency in PET performance characteristics.

## 2. Materials and Methods

### 2.1. Phantom Conceptualization and Fabrication

Cubic grids of 4 × 4 × 4 cm^3^ were created in Fusion 360 (Autodesk Inc, San Francisco, CA, USA) with the Volumetric Lattice toolset. The 4 cm size was chosen to accommodate the smallest to largest lesions (Eg. 4 mm to 38 mm) normally emulated in conventional practices **[4,6,7]**. Two essential design characteristics were incorporated: solidity (*S*) and pore distance (*D_P_*). Solidity is the percentage of solid material that regulates the fillable volume, establishing the contrast between the target and background, while pore distance is the distance between two consecutive pores (a.k.a. base unit cell) which dictates the permeability (ease of volumetric filling by the fluid) and the limit of the spatial resolution of the printed object. A key design challenge for the grid is to balance and achieve a suitable trade-off that enhances activity dispersion and the removal of air bubbles from the mesh, and accommodates the scanner spatial resolution. Three *S* configurations (50, 60, and 70%) and three *Dp* variants (5,7,10 mm) were investigated as shown in (**Fig. 1.a**).

**Figure 1.**
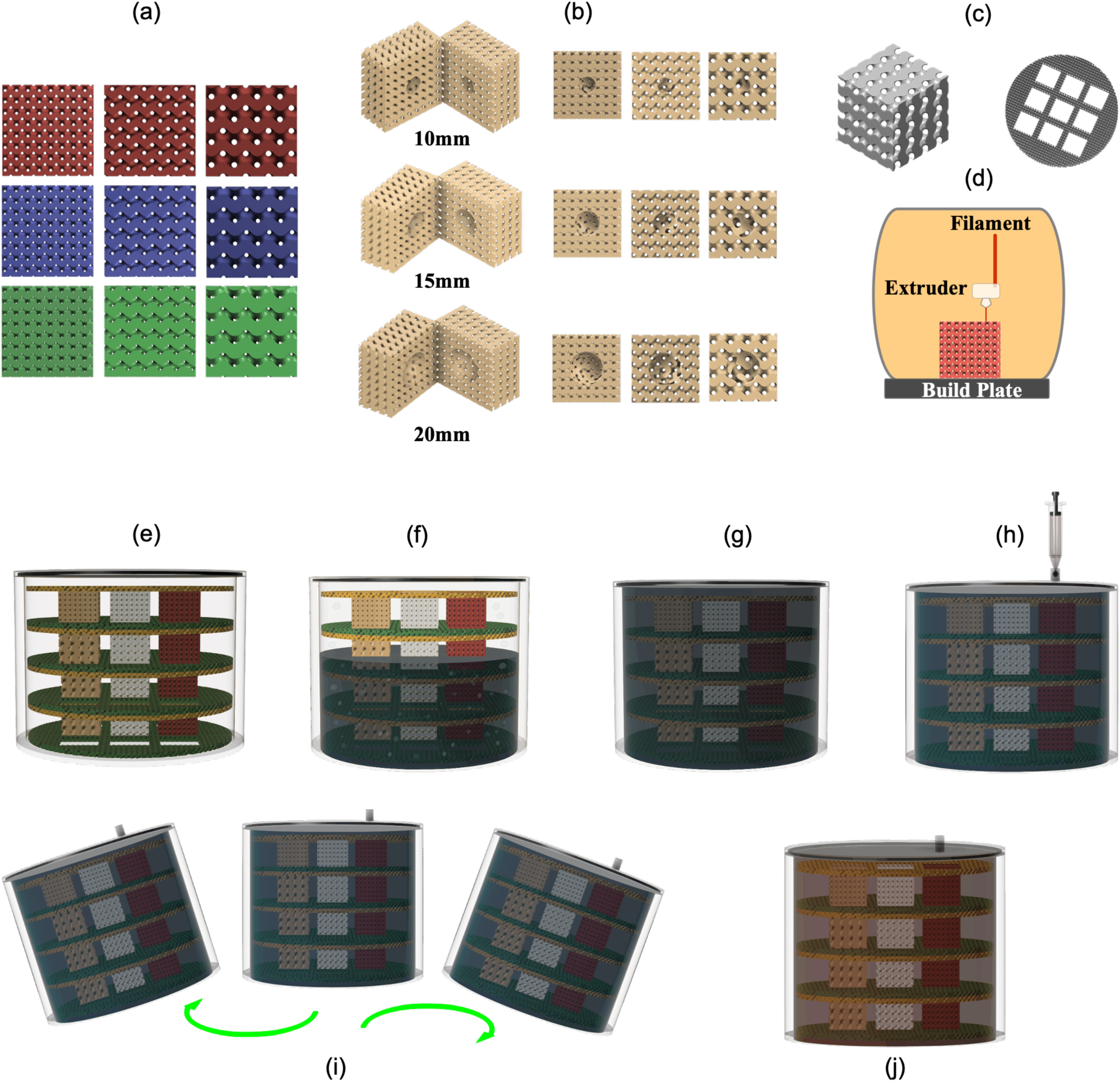
Schematic of grid design and fabrication: (**a**) Central section of baseline 6 design parameters, Rows - *S*: 50% (red), 60% (blue), 70% (green) (top to bottom) and Columns – *D_P_*: 5 mm, 7 mm, 10 mm (left to right); (**b**) Split cubes and central Slice for varying *D_P_*, showing different target voids (**c**) Rendering of cube insert and designed Jaszczak cube holder used for G-code generation; (**d**) FDM illustration for fabrication of the designed inserts; Phantom preparation protocol: (**e**) placement of support and cubes inside the phantom, (**f**) filling phantom with soap and water solution, (**g**) phantom relaxation, (**h**) injection of activity, (**i**) mixing post-injection by shaking, (**j**) radioactive phantom ready for imaging

The target voids of varying diameters (*T*: 10, 15, and 20 mm) were created by a Boolean subtraction centred on the cubic grids (**Fig. 1.b**) and the spherical shape was chosen based on conventional sphere sizes. These spherical voids are completely open, allowing them to be fully filled with the radioactive tracer solution, thereby reaching the intended activity concentration (AC) as :

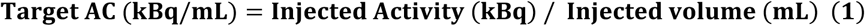

In contrast, the surrounding mesh contains alternating solid material, where the solid material contributes zero tracer theoretically, and interconnected pores. For a given solidity, the effective tracer concentration within the mesh is determined as:

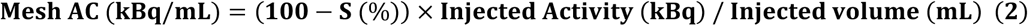

This difference in concentration makes the voids as “hotter” regions (emulating a lesion) against the dimmer background mesh (emulating tissue) on the acquired PET images. To streamline the design identification, *S-D_P_-T* nomenclature was used in order to be directly compatible with Fusion 360 as opposed to the fillable volume fraction (FVF) nomenclature previously used by **[9]**. A total of 36 cubes were designed, of which 9 were homogeneous cubes (no target) and the other cubes with spherical targets based on the possible permutations of the *S-D_P_-T*. Additionally, a meshed cube holder support (diameter 20 cm, thickness 7 mm, *S*=50 %, *D_p_* = 7 mm) with a 3 × 3 layout for cube insertion was designed to match the internal diameter of a commercial Jaszczak phantom cylinder as shown in (**Fig. 1.c**) in order to fix the cube’s position within the phantom. Gyroid patterns belonging to the TMPS (triply minimal periodic structures) family were used for the grid structure, having been found to have good directional fluid permeability compared to other patterns **[28]**. The models after meshing were exported as .STL files with a minimum resolution of 0.45 mm and were sliced for printing in a Prusa Slicer (Prusa Research, Prague, Czech Republic). Each set was then 3D-printed in ABS with the Prusa MK3S+ Printer (Prusa Research, Prague, Czech Republic). ABS is currently the optimal material to emulate water density **[27]**. The print parameters used can be found in **Table 1**. To characterize the printing accuracy, the theoretical expected weight for homogeneous cubes based on a 100% solidity cube were calculated. Printed density (*ρ*) was also calculated from measured weight and design volume (from Fusion 360). Five percent maximum deviation was deemed as an acceptable threshold for both weight and density.

**Table 1.**
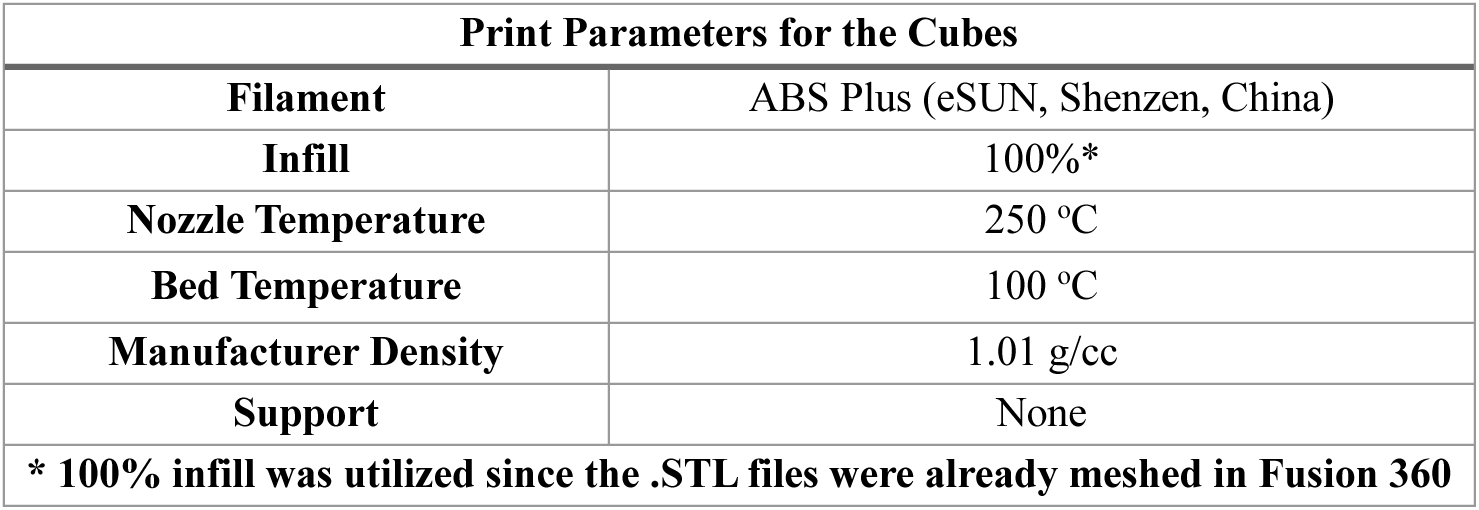
3D Printing Parameters used for each set.

| Print Parameters for the Cubes |  |
| --- | --- |
| **Filament** | ABS Plus (eSUN, Shenzhen, China) |
| **Infill** | 100%* |
| **Nozzle Temperature** | 250 °C |
| **Bed Temperature** | 100 °C |
| **Manufacturer Density** | 1.01 g/cc |
| **Support** | None |
| * 100% infill was utilized since the .STL files were already meshed in Fusion 360 |  |

Printed cubes were positioned within a commercial Jaszczak phantom cylinder. A simplified phantom preparation protocol is illustrated in **Fig 1. e-j**. Thirty-six cubes were arranged in 4 different layers of 9 per layer as show in in **Fig. 2. a-d**. The three main regions in the design were classified as radiotracer solution (no mesh), background mesh / grid, and targets of which an example is provided in **Fig. 2. e-f**. An *S* value of 50 %, 60 %, or 70 % corresponded to a TBR (target-to-background ratio) of 2, 2.5, and 3.33, respectively.

**Figure 2.**
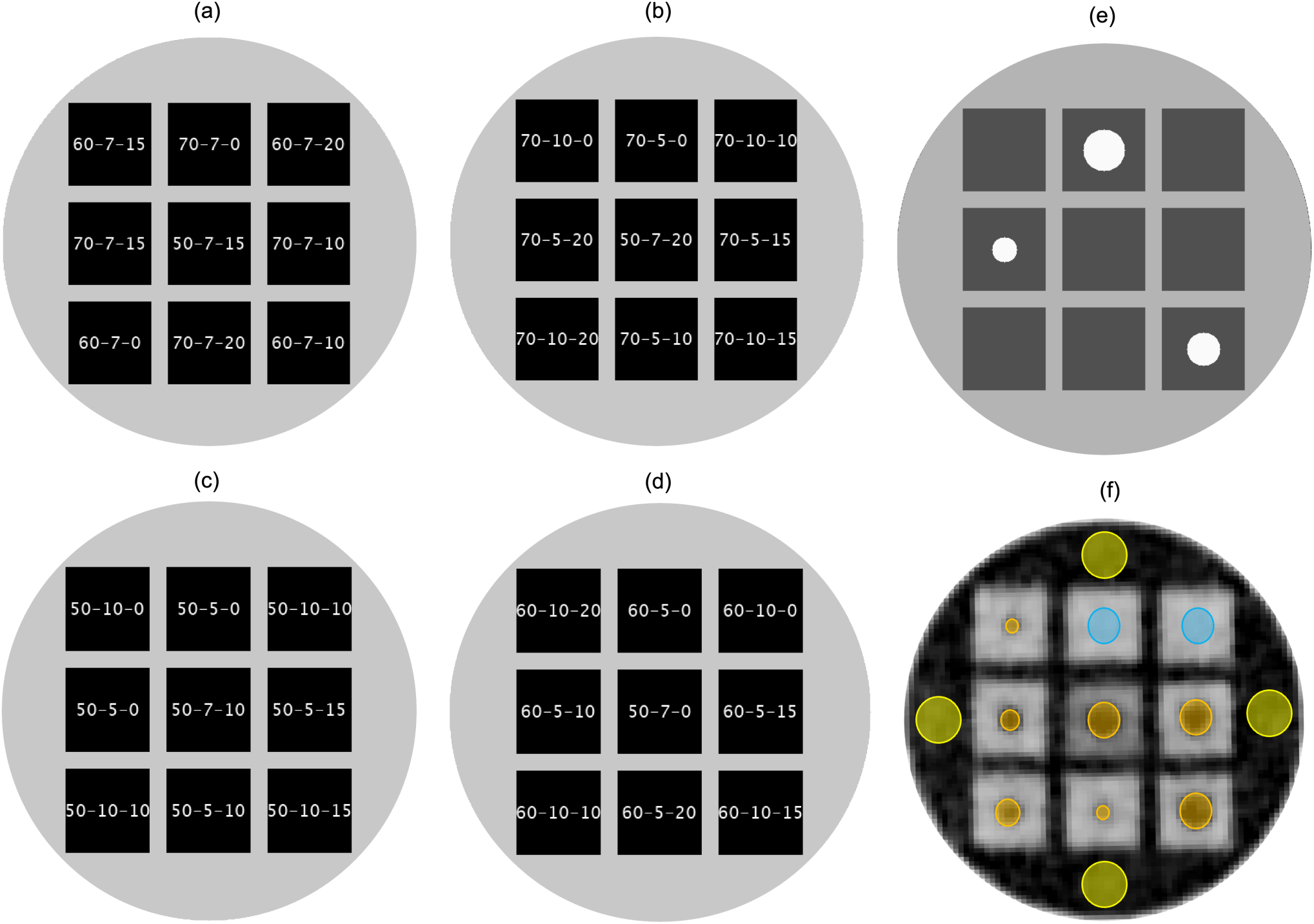
Layout of the 36 cubes inside the phantom subdivided into 4 layers – (**a**) Layer 1, (**b**) Layer 2, (**c**) Layer 3, (**d**) Layer 4; (**e**) Example illustration of different regions in the pet image – reference solution (light grey), background mimicking tissue (dark grey), target mimicking lesion (white) ; (**f**) VOI placement in different regions of each layer for data extraction – reference radiotracer solution (yellow), homogenous cubes (blue) and targets (orange)

Phantom filling was facilitated by a mixture of distilled water and soap, which was used to reduce the solution’s surface tension **[29]**. After the initial filling, the phantom was left for 6 hours with this soap solution to allow trapped air bubbles inside the grids to reach the top, beyond regions of interest. The phantom was filled only once with water (**Fig. 1. e-g**) on Thursday (Day 0) for the initial acquisition. Subsequent acquisitions followed on Friday (Day 1). It was left undisturbed over the weekend and used for three additional consecutive acquisitions from Monday to Wednesday (Days 4-6), totalling five acquisitions. The radiotracer was injected into the water (volume of 4.5 L) within the phantom. The tracer was administered using a single injection protocol, facilitating preparation compared to similar phantom studies—especially given the considerably increased number of targets. (**Fig. 2. h-j**). The tracer used in this study was [^18^F]FDG ([^18^F]-2-fluoro-2-deoxy-d-glucose), as per typical clinical protocols and was injected on each acquisition day as per **Table 2**.

**Table 2.** Injected Activity across different days.

| Acquisition Number | Injected Activity (MBq) | Decay Time (min) | Activity at Acquisition Start (MBq) | Day |
| --- | --- | --- | --- | --- |
| 1 | 82.8 | 60 | 56.1 | 0 |
| 2 | 66 | 73 | 41.2 | 1 |
| 3 | 64.3 | 45 | 47.5 | 4 |
| 4 | 60.4 | 30 | 47.5 | 5 |
| 5 | 54.4 | 25 | 45.4 | 6 |

### 2.2. Imaging and Reconstruction Parameters

All acquisitions were performed on a Discovery MI 5-ring digital PET-CT system (GE Healthcare, Chicago, IL, USA) with corrections and protocols similar to that of a clinical setting. CT images were acquired helically with slice thicknesses of 3.75 mm for CTAC, 2.5 mm for CT, and 1.25 mm for high resolution CT, with a pitch of 1.375, current of 89 mA, and peak voltage of 100 kV (noise index of 22). The CTAC interval was 2.8 mm to match the PET slice thickness. For each experimental iteration, 5 consecutive acquisitions of 5 minutes each centred on the phantom were performed. Major acquisition and reconstruction parameters used are summarized in **Table 3**.

**Table 3.** Acquisition Parameters.

| PET Acquisition Parameters |  |
| --- | --- |
| No. of Bed Positions | 1 |
| Matrix Size | 256 × 256 |
| Slice Thickness | 2.8 mm |
| No. of Slices | 89 |
| Scan Duration | 5 × 300 s |
| Reconstruction Diameter | 600 cm |
| Pixel Dimensions | 2.34 × 2.34 mm |
| Voxel Dimensions | 2.34 × 2.34 × 2.8 mm |
| Voxel Volume | 15.43 mm<sup>3</sup> |
| Reconstruction Algorithm | QCFX (time-of-flight BPL*) |
| Beta* | 550 |
| *Bayesian penalized-likelihood reconstruction with regularization parameter beta |  |

### 2.3. Data Extraction and Analysis

For qualitative assessment of this proof of concept, visual comparison of the transverse slices for the centre of each cube layer was performed for PET scans. The key features observed were the visual changes in the cubes across days, contrast across features and the visibility of the targets. Subsequently, quantitative data extraction was carried out using an AWServer client console with Volume Viewer, PETVCAR® (GE Healthcare, Chicagio, IL, USA) software similar to that of **[6]**. For each set of images, 27 spherical VOIs corresponding to the true internal volume of the targets were placed centered on the cubes. For the 9 homogenous cubes, VOI size was maintained same as the 20 mm target cubes. Additionally, 4 spherical VOIs were placed for each layer in reference solution adjacent to the cubes and were averaged per layer. The placement of VOI is shown in **Fig. 2.e**. AC (kBq/mL) for mean, maximum and standard deviation were extracted for all the VOI’s. The data extraction led to 1000 data points (900 in the cubes and 100 for the reference solution). The extracted PET data for each day was averaged over the respective 5 consecutive acquisitions thus leading to 180 insert VOI values (36 cubes × 5 days) and 20 reference values (4 layers × 5 days).

The SUV (standardized uptake value) was then calculated as below with a phantom weight of 8 kg decay corrected to the start of the PET acquisition:

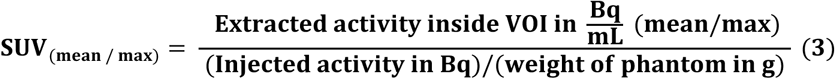

For quantitative analysis, two metrics were calculated from the SUV data: the target representation index (TRI) and dilution coefficient (DC). Both mean and maximum for these metrics were calculated using Equations 4 and 5:

- Target representation index represents the distribution and dispersion of activity concentration within the target voids relative to the reference solution around the printed cube.

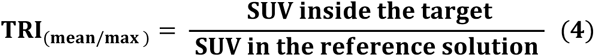

TRI is analogous to conventional RC (recovery coefficient) used in **[6]**. The key difference is that while RC quantifies the absolute signal recovery and loss due to the PVE (partial volume effect) w.r.t true value, TRI assesses the same relative to a reference volume measurement. This is because the exact “true” activity within each target isn’t independently injected unlike conventional procedure where the targets and background are filled separately. Therefore, the activity concentration in the large, well-mixed “reference volume”, insensitive to PVE serves as proxy for the expected “true” concentration that should be present *within* the voids. Also, this allows local evaluation for each of 4 layers in case of variability in activity distribution.

- Dilution coefficient represents the contrast between the printed homogeneous cube and the reference solution. This is same as contrast modulation coefficient mentioned in **[9]**, which confirms the percentage of tracer dilution to replicate tissues and is only used for homogeneous cubes in this work. Normally, DC_theoretical_ = 1 – (S/100)

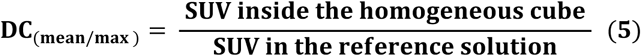

To assess the mixing of the radiotracer, SUV values of reference regions across 4 layers for each day were analyzed and a pairwise variability across layers were calculated. An acceptable threshold of 10% was chosen, similar to uniformity studies **[4,5]**. A subsequent statistical analysis on the dataset of PET metrics (TRI_mean, max_, DC_mean, max_) with N_sample_ : 180 for each was performed for normality through the Shapiro-Wilk test **[30]** separately on both mean and maximal values. The normality tests were complemented by a pairwise counterpart, Friedman Test **[31]** to assess global consistency across the metrics over 5 days. This test was chosen as its less sensitive to the results of normality tests. All statistical analyses were conducted in Integrated Development Environment for R (RStudio PBC, Boston, MA) with a predetermined significance level of *p <* 0.05.

## 3. Results

### 3.1. Assessment of Printing Accuracy

The 3D-printed cubes had high accuracy in the aspect of weight, with all measurements within 5% of expected values. The weight measurements of the cubes on the print day can be found in **Table 4**. The highest deviation of 3.125%, was observed for the 50-10-0 (*S*-*D_P_*-*T*) and 50-7-0 cubes. The density of the printed section of the grids was estimated to be between 0.97 – 1.03 g/mL, which is within 3 % of that of water (reference value: 1.00 g/mL). **Fig. 3. a** illustrates the printing of the cubes.

**Figure 3.**
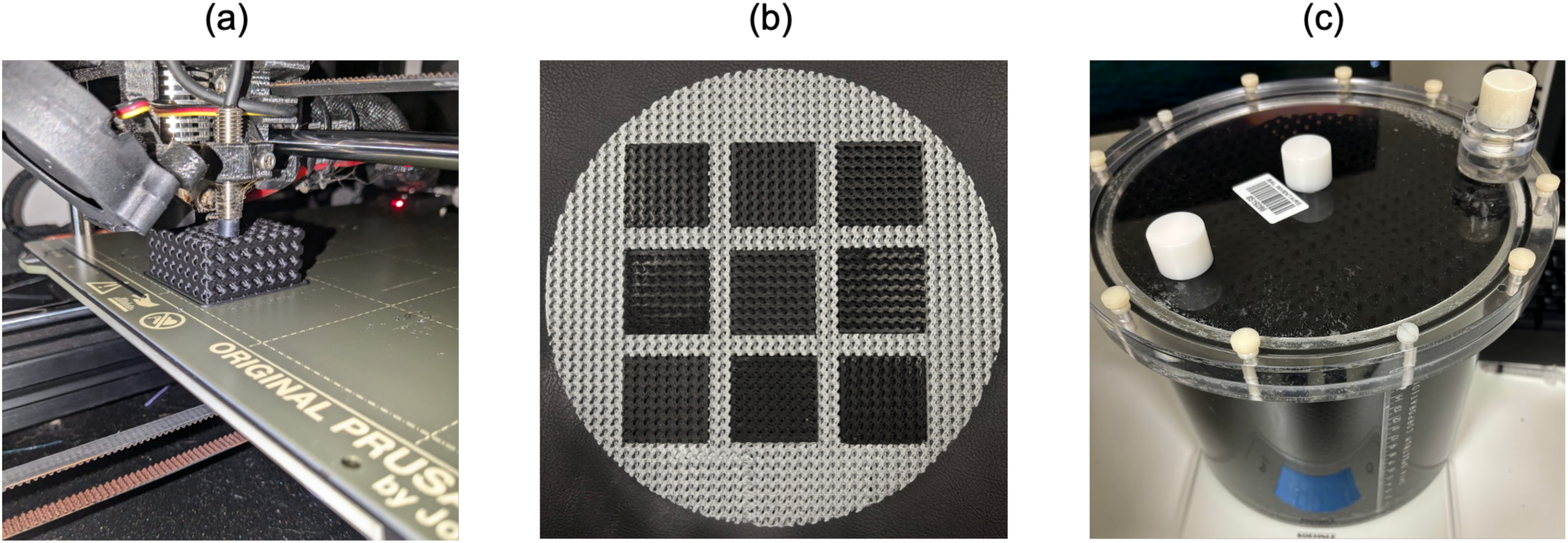
(a) Snapshot of cubes during printing; (b) Printed inserts arranged in a 3 × 3 layout using the cube holder; (c) Assembled Jaszczak phantom post-filling

**Table 4.** Measured Weight of 3D-Printed Homogeneous Cubes.

| Cube<br>(S-D-T) | Measured Weight<br>(g) | Expected Weight (g) | Difference in<br>Weight (%) | Effective Density<br>(g/mL) |
| --- | --- | --- | --- | --- |
| 50-5-0 | 32 | 32 | 0 | 1.00 |
| 50-7-0 | 33 | 32 | -3.125 | 1.03 |
| 50-10-0 | 31 | 32 | 3.125 | 0.97 |
| 60-5-0 | 37.5 | 38.4 | 2.34 | 0.98 |
| 60-7-0 | 37.3 | 38.4 | 2.86 | 0.97 |
| 60-10-0 | 38 | 38.4 | 1.04 | 0.99 |
| 70-5-0 | 44 | 44.8 | 1.79 | 0.98 |
| 70-7-0 | 44 | 44.8 | 1.79 | 0.98 |
| 70-10-0 | 44.1 | 44.8 | 1.56 | 0.98 |

### 3.2. PET Uptake across Five Days

Contrast differences in the PET images viewed with the same SUV display window (0-2 g/mL) are illustrated in **Fig 4.a**. Clear demarcation is visible among contrasts between the background grid, target voids, and the reference radiotracer solution. The targets are all distinguishable, but less so as the size decreases. The 70% solidity cubes are the most distinct, and the 50% solidity cubes are the least distinct in terms of visual contrast. Also visually evident is that the activity concentration in the background solution varies between different layers of the phantom. The SUV_mean_ and SUV_max_ of the reference solution are presented in **Fig 4.b, 4.c** and the corresponding variability test results can be found in the **Supplementary Information (Fig. S3, S4).** DC values for the 9 homogeneous cubes are presented in **Fig 5.a**. DC expected for respective solidities were 0.5, 0.4, and 0.3. For 50 % solidity, the DC values fluctuated around the theoretical value of 0.5, with extreme outliers based on the pore distance and days. For 60 % solidity, the coefficients of dilution ranged from 0.35 to 0.48, and for 70 % solidity, the dilution coefficients were consistently higher than expected, with values ranging from 0.32 to 0.40. TRI values as function of target size are presented in **Fig**. **5****.b-d**. TRI values varied over the five acquisition days for all three pore distances 5 mm, 7 mm, and 10 mm but no clear quantitative trend was seen as function of acquisition day. The larger spherical targets (20 mm) consistently demonstrated TRI values closer to ideal expectations in contrast to the results for the 10 mm targets. The results of statistical tests can be found in **Table 5**. All the data was found to be non – normal based on the Shapiro-Wilk Normality Test. Friedman tests for TRI and DC are also included. Additionally, results of a post-hoc Wilcoxon test with Bonferroni adjustment was also performed and corresponding results are available in the **Supplementary Information (Fig. S5)** for more statistical inference.

**Figure 4.**
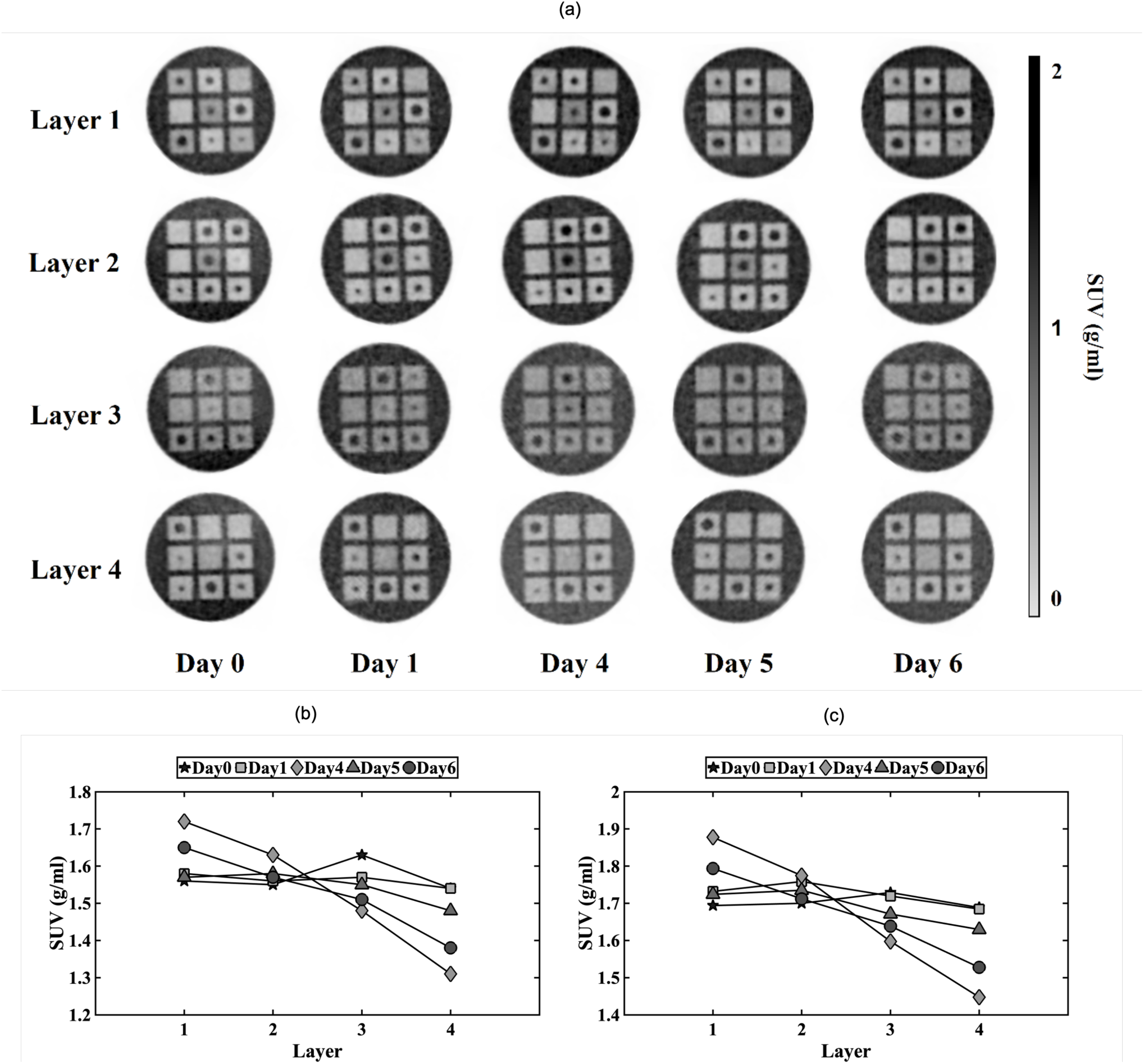
(a) Central axial PET slice for each layer over 5 acquisition days viewed on same SUV window scale; SUV variation across 4 layers (b) SUV_mean_ (c) SUV_max_;

**Figure 5.**
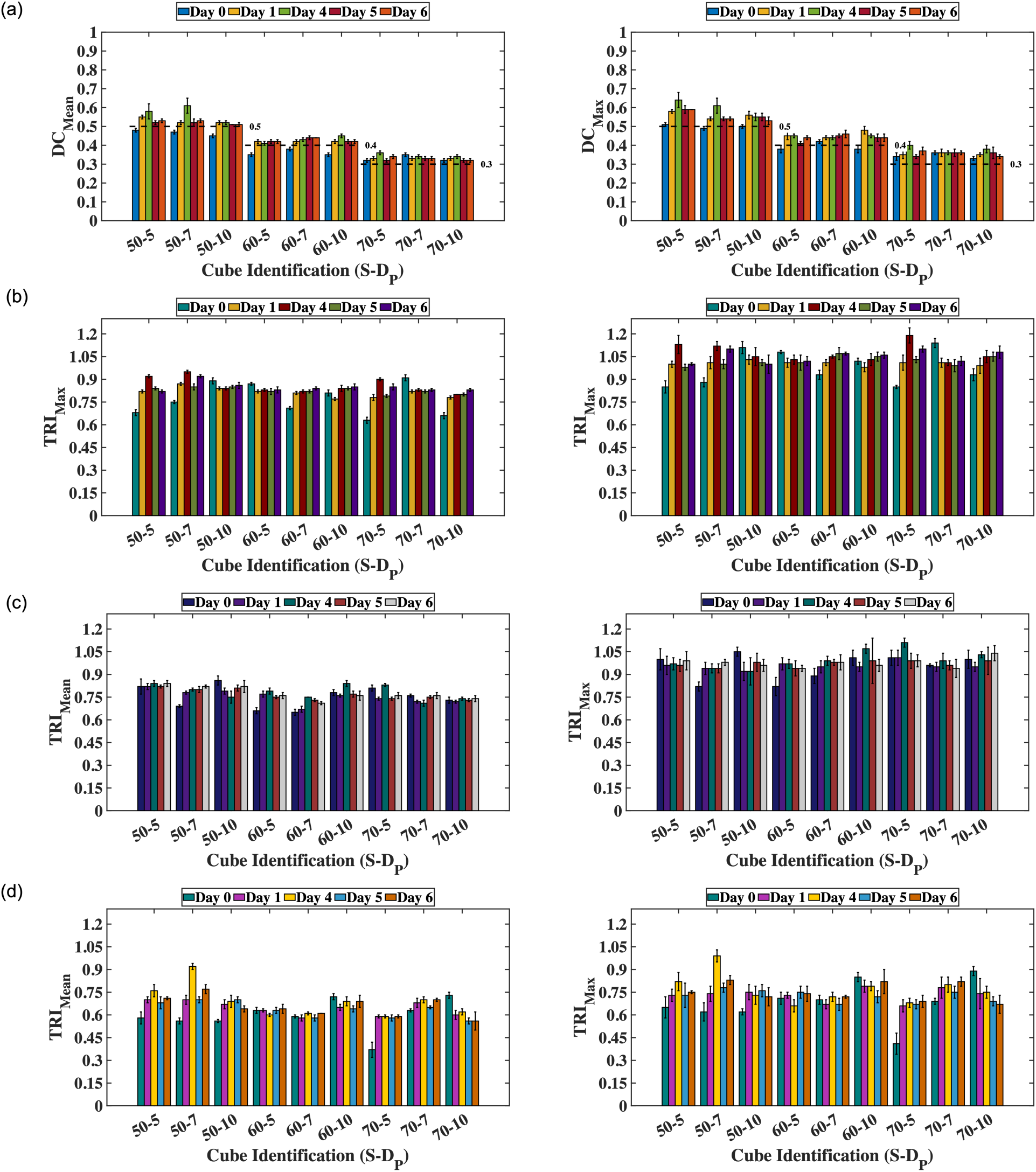
Quantitative metrics calculated with mean values (left), maximum values (right) - (a) DC for homogeneous cubes, TRI for a target size of (b) 20 mm, (c) 15 mm, (d) 10 mm ; Error bars represented are standard deviation – The same values can be found in tabular form (**Supplementary Information tables S1, S2, S3, S4**)

**Table 5.** Summary of Statistical Tests.

| Statistical Test | Key Results (Significance – 0.05) |
| --- | --- |
| Shapiro-Wilk Normality Test | TRI<sub>max</sub>: $W = 0.91, p\text{-value} < 0.01$<br>TRI<sub>mean</sub>: $W = 0.93, p\text{-value} < 0.01$<br>DC<sub>max</sub>: $W = 0.93, p\text{-value} < 0.01$<br>DC<sub>mean</sub>: $W = 0.91, p\text{-value} < 0.01$ |
| Friedman Test | TRI<sub>max</sub>: $p\text{-value} = 0.67$; TRI<sub>Mean</sub>: $p\text{-value} = 0.93$<br>DC<sub>max</sub>: $p\text{-value} = 0.03$; DC<sub>Mean</sub>: $p\text{-value} = 0.07$ |

## 4. Discussion

Low cost FDM based 3D-printed cubic inserts were explored to create targets in PET/CT quality control (QC) phantoms. Printing accuracy of the cubes weighing between ± 5% of expectation indicates the accuracy of FDM in the production of phantom inserts with a precision crucial for the true representation of the planned experimental setup. These also fall within the prescribed thresholds of previous 3D printing-based studies **[8,9,32]**. The equivalent density of the solid component (0.97 – 1.03 g/cc) falls within the range of water **[27]** which is a preferred property for emulating the high energy photon attenuation of tissue.

Evident visible contrast variations in the PET images verified that three regions could be distinguished: the background grid, target voids, and reference radiotracer solution. This is essential for the accurate quantification of radiotracer uptake. The fact that the 70 % solidity cubes had the maximum visual contrast, whereas 50% solidity cubes had the minimum visible contrast, highlights that the intended concept of different contrasts through a single injection of tracer is feasible and reproducible with FDM. Additionally, as the target size becomes smaller, the visibility becomes poorer due to PVE, consistent with conventional findings.

The proposed protocol in this study requires single injection of activity in a prefilled phantom rather than a stock prediluted strategy used previously by **[9,18]**, a key advantage being the method’s lower susceptibility to air bubbles, which has been an issue in previous studies. Because of its prolonged immersion strategy, the proposed protocol had no air bubbles inside target voids at the scanning spatial resolution of the PET (**Fig. 4a)** or CT (**Supplementary information, Fig S1**). However, relying on a single injection of activity can increase the susceptibility to mixing discrepancies causing a SUV distribution variability along the axis of the phantom, in this case. Non-uniformity of the activity dispersion in background across phantom layers was apparent as seen in **Fig. 4a**. The layer variability for Day-4 and Day-6 values fail the 10% variability threshold, as shown in the **Supplementary Information (Fig. S3, S4)** with maximum variability reaching 30%. This is also supported by the post-hoc follow up test **Supplementary Information (Fig. S5),** where the comparisons lead to statistically significant differences for Day-4 and Day-6 pairs. However, with careful mixing, the acceptable standard for such studies could be maintained as 10% variability across layers. For future studies, removing volume of background solution (example 50 mL) close to the filling cap of the phantom prior to the injection and adding it back prior to scan could be explored to reduce the layer-to-layer variation **[5]**.

In the quantitative metrics, the target representation index was consistent across days supported by Freidman tests p-values (**Table 5**). One pattern that we observe in **Fig. 4.a and 5.b-d** is that no specific *D_p_* effects the target representation particularly confirming that the pores were well under the PET spatial resolution similar to that of findings in **[9]**. For a *D_P_* of 5 mm, 69% of TRI _max_ values and 39% of TRI _mean_ values yielded greater than 80% recovery. For a *D_P_* of 7 mm, 62% of the TRI _max_ values and 32% of the TRI _mean_ values exceeded same threshold. For a *D_P_* of 10 mm, the distribution observed was 65% for TRI _max_ and 36% for the TRI _mean_. This indicates that the pore distance, within the range tested, did not have a significant influence on the overall tracer recovery. This is consistent with previous studies, in that the fillable volume fraction (solidity) of the printed inserts is the key factor driving PET performance, rather than pore distance **[24]**. Interestingly, individual DC_mean_ and DC_max_ values (**Fig. 5.a**) indicated a slight systematic bias in measured dilution compared to theoretical values. This is also confirmed by the p-values of Friedman test being very close to 0.05 (for mean) and < 0.05 (for max). DC values of 50% cubes from Day 4 had extreme outliers (**Fig. 5.a**) due to non-homogenous mixing which led to statistically differences. We performed a retest excluding Day - 4 DC, which resulted in Friedman *p* values of (DC_max_: 0.27) and (DC_mean_: 0.11). Of the two metrics, TRI is less sensitive to this non uniformity in mixing due to its relative nature. Although globally the achieved dilution is within the thresholds of 10%, the precise value for each experiment can vary. Even **[9]** reported the relation between dilution and fillable volume as linear with an intercept denoting the bias. This is partially due to printing inaccuracies but mostly a function of noise especially in case of DC_Max_.

Interestingly, the CT visibility depends on S and D_P_ strongly in contrast to PET behavior. For instance, the HU values of D_P_ = 5 mm change from −21.4±4.9 to −4.4±3.7 (for S: 50), −21.9±3.5 to −8.7±3.7 (for S: 60) and −28.4±4.9 to −14.4±3.5 (for S: 70) over the immersion period of 7 days. This drift towards HU of water is due to improved structural integrity of the ABS cubes in which air spaces caused by printing inaccuracies are filled by water due to prolonged immersion. Ideally, this HU drift should stabilize once it reaches very close to water as there would be no more air pockets to be filled **[33,34]**. The same was observed when the D_P_ = 5 mm cubes were tested after 6 months of immersion, where the mean values were between 7.6 ± 4.3 to 4.5 ± 3.5 falling within range of water (3.1 ± 3.7). The CT attenuation observed was in the range of - 28 to -5 which is significantly closer to water than the range of 90-150 **[8,9,23]** observed in previous studies. Also, D_P_: 7, 10mm resulted in grid resolution higher than CT spatial resolution, thus explicit visibility on CT images **(Supplementary information Fig. S1, Table. S1)** despite prolonged immersion. But importantly, no evidence on impact of this HU drift on PET AC correction was found in this study. Though not relevant within this framework, for studies aiming to benchmark the current design for quality control (QC) purposes where the material properties need to be standardized, *D_P_* should be optimized further for CT resolution and water attenuation i.e. immersion history needs to be taken into account until the HU values stabilize.

The most significant finding in this study is that 36 targets can be analyzed at once rather than 6 fixed targets in a conventional experimental setup such as **[6]**. In terms of impact, this could be reflected as 6 times fewer experiments for the same target sizes and a corresponding reduction in radiation exposure during phantom loading. The compatibility of this phantom design with conventional PET metrics such as SBR (signal-to-background ratio) or CNR (contrast-to-noise ratio) was not performed within the framework of this study as the main goal was evaluation of design parameters and consistency of chosen metrics rather than benchmarking against conventional phantoms. Also, for such an evaluation, a more robust automatic data extraction method is required over the manual method used in this work.

A significant contribution of this study is the modest cost of the inserts; each insert and cube holder on average cost less than 5€ (estimated by Prusa slicer). Additionally, other costs such as the printer, material, electricity consumption which would total to an overall cost estimation of less than 1000€ which is considerably lower than polyjet. These costs are similar to that of SLA presented by **[9,23]** but with improved CT attenuation (closer to that of water). Another major advantage for grid-based studies are their inherent capability to also model heterogeneity, for example by creating a second grid within the target void. Besides this, anthropomorphic lesions, which are directly segmented out of patient scans, can be translated into grid-based insert designs, effectively simulating patient-specific anatomy and activity distributions. A threshold for *D_P_* that could account for fluid permeability and scanner spatial resolution across different S values to optimize quantitative performance is to be established. With the development of more comprehensive datasets for a wider range of design parameters and printing conditions, limitations discussed in this work can be addressed. Therefore, we conclude that FDM based ABS inserts for PET produce consistent results which are both reliable and reproducible whilst comparable to that of SLA grids and can be extended for more anthropomorphic studies.

## 5. Conclusion

Feasibility of FDM based 3D-printed inserts was established for PET/CT applications. The quantitative metrics were found to be consistent globally across several acquisitions. The main contributions from this study are as follows

1. Simplified phantom preparation protocol (single injection) with attendant reduction in radiation exposure
2. Increased control on design of features (customizability)
3. Increasing the number of targets scanned per experiment
4. Reduced costs of 3D-printed inserts

## Abbreviations

ABS: Acrylonitrile Butadiene Styrene
AC: Activity Concentration in kBq/ml
CNR: Contrast to Boise Ratio
DC: Dilution Coefficient
DMI: Discovery MI
D_P_: Distance between pores
FDM: Fused Deposition Modelling
FDG: Fluorodeoxyglucose
FOV: Field of View
HU: Hounsfield Units
IEC: International Electrotechnical Commission
NEMA: National Electrical Manufacturing Association
NM: Nuclear Medicine
PET / CT: Positron Emission Tomography / Computed Tomography
PLA: Polylactic Acid
QC: Quality Control
RC: Recovery Coefficient
S: Solidity
SBR: Signal to Background Ratio
SD: Standard Deviation
SLA: Stereolithography
SPECT/CT: Single Photon Emission Computed Tomography / Computed Tomography
SUV: Standardized Uptake Value
TOF: Time of Flight
TBR: Target to Background Ratio
TMPS: Triply Minimal Periodic Structures
TRI: Target Representation Index
VOI: Volume of Interest

## Data Availability

All data produced in the present study are available upon reasonable request to the authors

## Supplementary Information

### CT Images and HU values

This section provides the central cross-sectional images of corresponding CT images used for attenuation correction of PET images presented in the main article.

**Figure S1.**
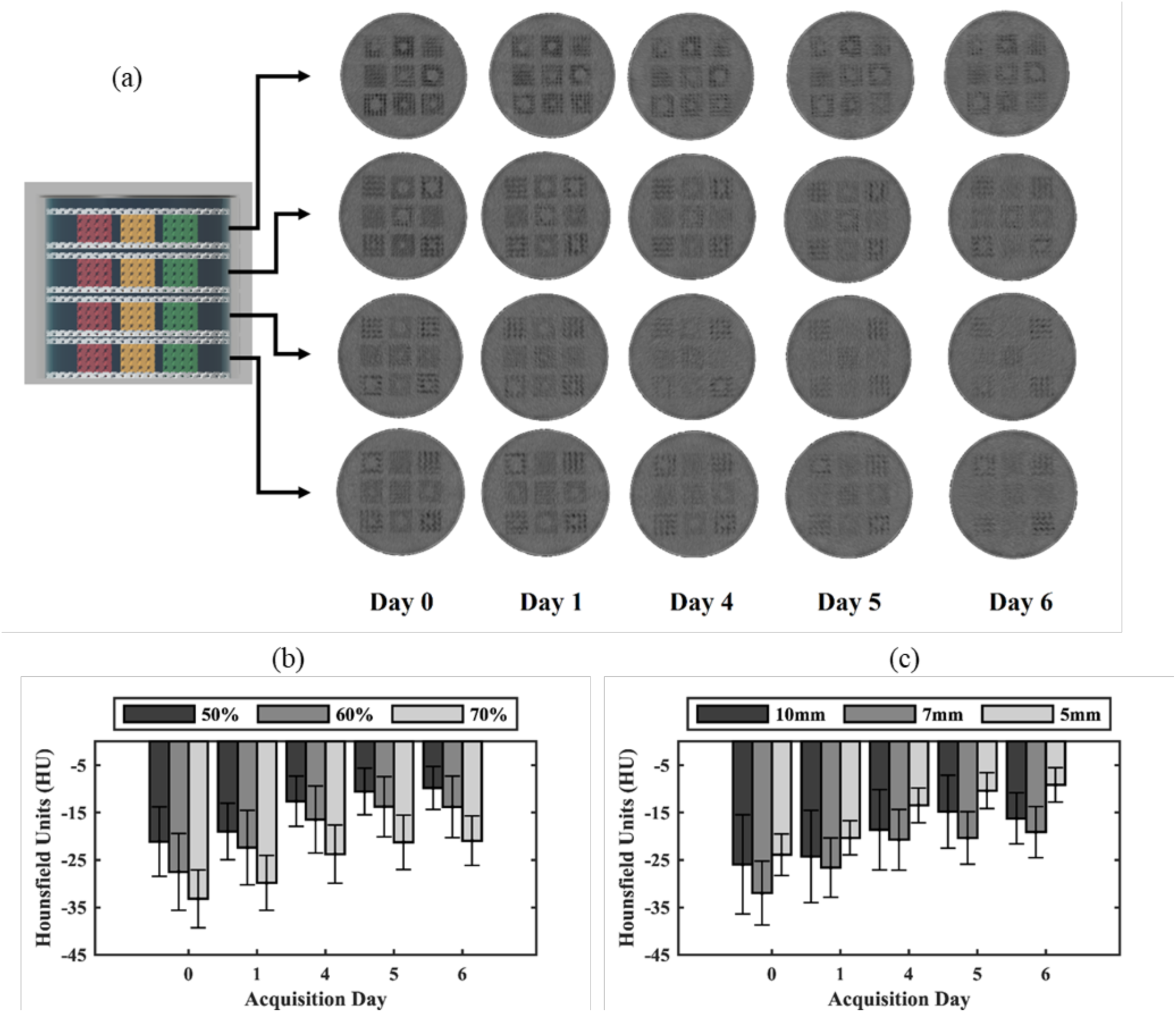
(a) CT progression of central axial slice over 5 acquisition days viewed on abdominal windowing; (b); Averaged HU_mean_ values of homogeneous cubes as function of acquisition day and S (c) Averaged HU_mean_ values of homogeneous cubes as function of *D_P_* and acquisition day

**Figure S2.**
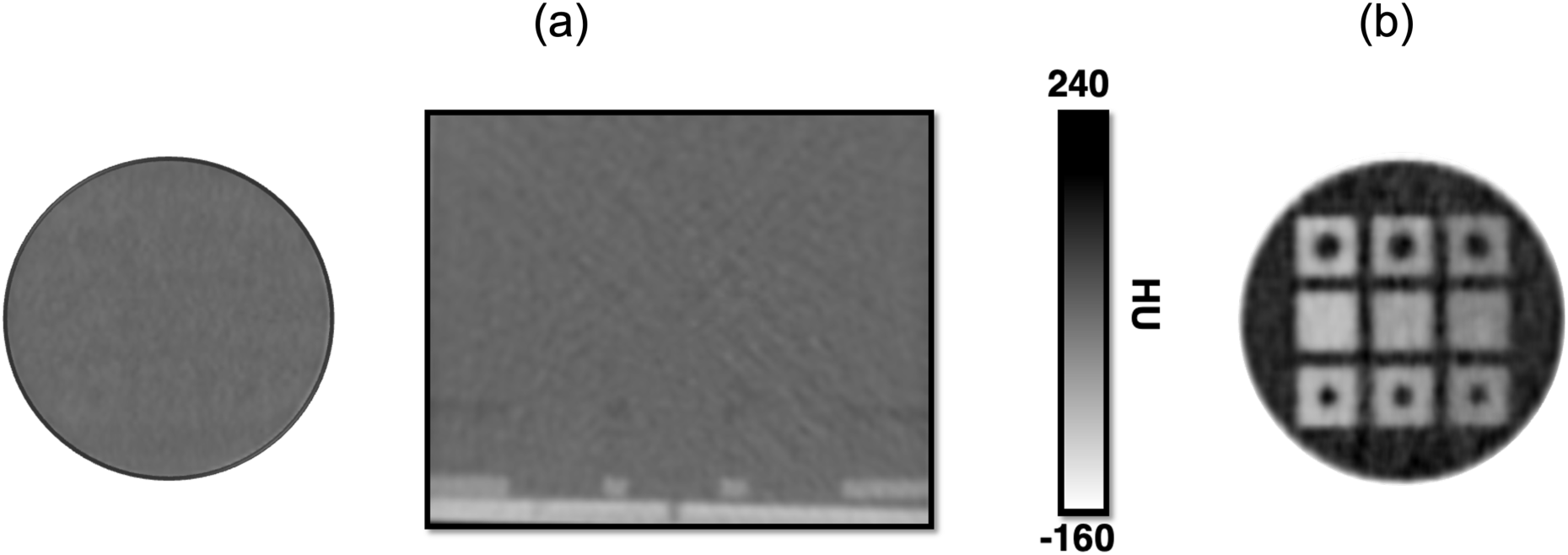
(a) CT & (b) PET Images of the D_p_ : 5mm cubes after 6 months of immersion carried out for a different study with layout S: 70, 60, 50 % (Columns) & T: 20, 0, 15mm (Rows). Results added for illustration that CT stabilises over the immersion time.

### Quantitative Metrics

This section provides the data plotted in the main article in tabulated form. Additionally the pairwise layer variability for reference solution is also presented to signify the non-uniform mixing discussed in the main article.

**Table S1.** Dilution Coefficient values for Homogeneous Cubes.

| Solidity % | Pore Distance mm | Dilution Coefficient (Max ± SD & Mean ± SD) | | | | |
| --- | --- | --- | --- | --- | --- | --- |
|  |  | Day 0 | Day 1 | Day 4 | Day 5 | Day 6 |
| 50 | 5 | 0.51±0.01 | 0.58±0.01 | 0.64±0.04 | 0.59±0.02 | 0.59±0.00 |
| | | 0.48±0.01 | 0.55±0.01 | 0.58±0.01 | 0.52±0.01 | 0.53±0.00 |
| | 7 | 0.49±0.01 | 0.54±0.01 | 0.67±0.04 | 0.54±0.01 | 0.54±0.01 |
| | | 0.47±0.01 | 0.52±0.01 | 0.61±0.04 | 0.52±0.02 | 0.53±0.01 |
| | 10 | 0.50±0.01 | 0.56±0.02 | 0.55±0.02 | 0.55±0.02 | 0.53±0.02 |
| | | 0.45±0.01 | 0.52±0.01 | 0.52±0.01 | 0.51±0.00 | 0.51±0.01 |
| 60 | 5 | 0.38±0.02 | 0.45±0.02 | 0.45±0.01 | 0.44±0.01 | 0.44±0.01 |
| | | 0.35±0.01 | 0.41±0.01 | 0.41±0.01 | 0.42±0.01 | 0.42±0.01 |
| | 7 | 0.42±0.01 | 0.44±0.01 | 0.44±0.01 | 0.45±0.01 | 0.46±0.02 |
| | | 0.38±0.01 | 0.42±0.01 | 0.43±0.01 | 0.44±0.01 | 0.44±0.00 |
| | 10 | 0.38±0.02 | 0.48±0.02 | 0.48±0.02 | 0.44±0.02 | 0.44±0.02 |
| | | 0.35±0.01 | 0.42±0.01 | 0.45±0.01 | 0.42±0.01 | 0.42±0.01 |
| 70 | 5 | 0.34±0.02 | 0.35±0.02 | 0.40±0.02 | 0.34±0.01 | 0.37±0.02 |
| | | 0.32±0.01 | 0.33±0.01 | 0.36±0.01 | 0.32±0.01 | 0.34±0.01 |
| | 7 | 0.36±0.01 | 0.36±0.02 | 0.36±0.01 | 0.36±0.02 | 0.36±0.01 |
| | | 0.35±0.01 | 0.33±0.01 | 0.34±0.01 | 0.33±0.01 | 0.33±0.01 |
| | 10 | 0.33±0.01 | 0.35±0.01 | 0.38±0.02 | 0.36±0.03 | 0.34±0.01 |
| | | 0.32±0.01 | 0.33±0.01 | 0.34±0.01 | 0.32±0.01 | 0.32±0.01 |

**Table S2.**
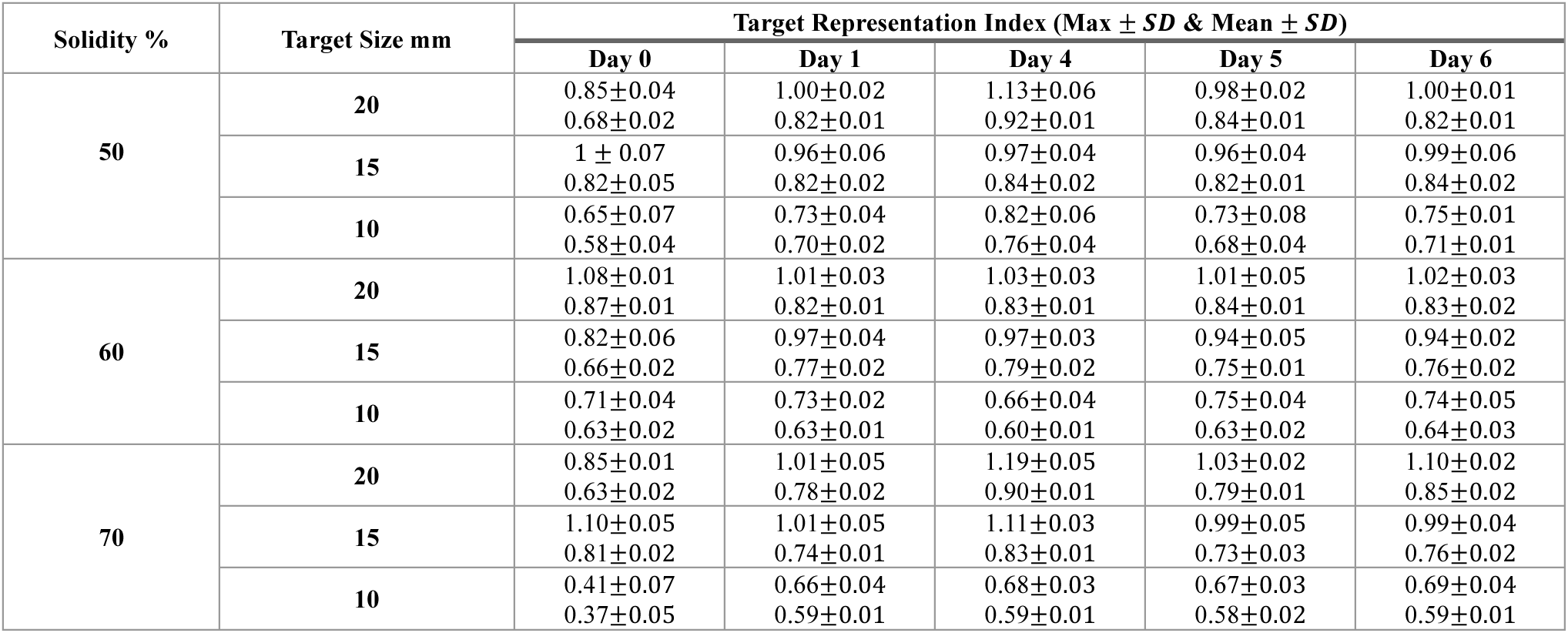
Target Representation Index values for Target Cubes (D_P_: 5mm)

| Solidity % | Target Size mm | Target Representation Index (Max ± SD & Mean ± SD) | | | | |
| --- | --- | --- | --- | --- | --- | --- |
|  |  | Day 0 | Day 1 | Day 4 | Day 5 | Day 6 |
| 50 | 20 | 0.85±0.04 | 1.00±0.02 | 1.13±0.06 | 0.98±0.02 | 1.00±0.01 |
| | | 0.68±0.02 | 0.82±0.01 | 0.92±0.01 | 0.84±0.01 | 0.82±0.01 |
| | 15 | 1 ± 0.07 | 0.96±0.06 | 0.97±0.04 | 0.96±0.04 | 0.99±0.06 |
| | | 0.82±0.05 | 0.82±0.02 | 0.84±0.02 | 0.82±0.01 | 0.84±0.02 |
| | 10 | 0.65±0.07 | 0.73±0.04 | 0.82±0.06 | 0.73±0.08 | 0.75±0.01 |
| | | 0.58±0.04 | 0.70±0.02 | 0.76±0.04 | 0.68±0.04 | 0.71±0.01 |
| 60 | 20 | 1.08±0.01 | 1.01±0.03 | 1.03±0.03 | 1.01±0.05 | 1.02±0.03 |
| | | 0.87±0.01 | 0.82±0.01 | 0.83±0.01 | 0.84±0.01 | 0.83±0.02 |
| | 15 | 0.82±0.06 | 0.97±0.04 | 0.97±0.03 | 0.94±0.05 | 0.94±0.02 |
| | | 0.66±0.02 | 0.77±0.02 | 0.79±0.02 | 0.75±0.01 | 0.76±0.02 |
| | 10 | 0.71±0.04 | 0.73±0.02 | 0.66±0.04 | 0.75±0.04 | 0.74±0.05 |
| | | 0.63±0.02 | 0.63±0.01 | 0.60±0.01 | 0.63±0.02 | 0.64±0.03 |
| 70 | 20 | 0.85±0.01 | 1.01±0.05 | 1.19±0.05 | 1.03±0.02 | 1.10±0.02 |
| | | 0.63±0.02 | 0.78±0.02 | 0.90±0.01 | 0.79±0.01 | 0.85±0.02 |
| | 15 | 1.10±0.05 | 1.01±0.05 | 1.11±0.03 | 0.99±0.05 | 0.99±0.04 |
| | | 0.81±0.02 | 0.74±0.01 | 0.83±0.01 | 0.73±0.03 | 0.76±0.02 |
| | 10 | 0.41±0.07 | 0.66±0.04 | 0.68±0.03 | 0.67±0.03 | 0.69±0.04 |
| | | 0.37±0.05 | 0.59±0.01 | 0.59±0.01 | 0.58±0.02 | 0.59±0.01 |

**Table S3.** Target Representation Index values for Target Cubes (D_P_: 7 mm)

| Solidity % | Target Size mm | Target Representation Index (Max ± SD & Mean ± SD) | | | | |
| --- | --- | --- | --- | --- | --- | --- |
|  |  | Day 0 | Day 1 | Day 4 | Day 5 | Day 6 |
| 50 | 20 | 0.88±0.03 | 1.01±0.04 | 1.12±0.03 | 1 ± 0.03 | 1.10±0.02 |
| | | 0.75±0.01 | 0.87±0.01 | 0.95±0.01 | 0.85±0.02 | 0.92±0.01 |
| | 15 | 0.82±0.07 | 0.94±0.03 | 0.94±0.05 | 0.94±0.03 | 0.98±0.03 |
| | | 0.69±0.03 | 0.78±0.01 | 0.80±0.01 | 0.80±0.01 | 0.82±0.02 |
| | 10 | 0.62±0.06 | 0.74±0.05 | 0.99±0.04 | 0.78±0.03 | 0.83±0.03 |
| | | 0.56±0.02 | 0.70±0.03 | 0.92±0.02 | 0.70±0.02 | 0.77±0.03 |
| 60 | 20 | 0.93±0.03 | 1.01±0.02 | 1.05±0.01 | 1.07±0.04 | 1.07±0.01 |
| | | 0.71±0.01 | 0.81±0.01 | 0.82±0.01 | 0.82±0.01 | 0.84±0.01 |
| | 15 | 0.89±0.05 | 0.95±0.04 | 0.99±0.03 | 0.98±0.02 | 0.98±0.05 |
| | | 0.65±0.02 | 0.67±0.02 | 0.71±0.00 | 0.73±0.01 | 0.71±0.01 |
| | 10 | 0.70±0.03 | 0.67±0.03 | 0.72±0.03 | 0.67±0.04 | 0.72±0.01 |
| | | 0.59±0.01 | 0.58±0.02 | 0.61±0.01 | 0.58±0.02 | 0.61±0.00 |
| 70 | 20 | 1.14±0.03 | 1.01±0.03 | 1.01±0.02 | 0.99±0.04 | 1.02±0.03 |
| | | 0.91±0.02 | 0.82±0.01 | 0.83±0.01 | 0.82±0.01 | 0.83±0.01 |
| | 15 | 0.96±0.01 | 0.95±0.03 | 0.99±0.05 | 0.96±0.03 | 0.94±0.06 |
| | | 0.76±0.01 | 0.72±0.01 | 0.71±0.02 | 0.75±0.01 | 0.76±0.02 |
| | 10 | 0.69±0.02 | 0.78±0.01 | 0.80±0.05 | 0.75±0.04 | 0.82±0.03 |
| | | 0.63±0.01 | 0.68±0.03 | 0.70±0.02 | 0.65±0.01 | 0.70±0.01 |

**Table S4.** Target Representation Index values for Target Cubes (*D_P_*: 10mm)

| Solidity % | Target Size mm | Target Representation Index (Max ± SD & Mean ± SD) | | | | |
| --- | --- | --- | --- | --- | --- | --- |
|  |  | Day 0 | Day 1 | Day 4 | Day 5 | Day 6 |
| 50 | 20 | 1.11±0.04 | 1.03±0.03 | 1.05±0.06 | 1.01±0.02 | 1.04±0.06 |
| | | 0.89±0.02 | 0.84±0.01 | 0.84±0.01 | 0.85±0.01 | 0.86±0.02 |
| | 15 | 1.05±0.02 | 0.92±0.06 | 0.92±0.09 | 0.98±0.06 | 0.96±0.04 |
| | | 0.86±0.03 | 0.79±0.02 | 0.75±0.04 | 0.81±0.02 | 0.82±0.02 |
| | 10 | 0.62±0.02 | 0.75±0.05 | 0.73±0.06 | 0.76±0.04 | 0.72±0.06 |
| | | 0.56±0.01 | 0.67±0.03 | 0.69±0.04 | 0.70±0.02 | 0.64±0.02 |
| 60 | 20 | 1.02±0.02 | 0.98±0.03 | 1.03±0.04 | 1.05±0.03 | 1.06±0.02 |
| | | 0.81±0.02 | 0.77±0.01 | 0.84±0.02 | 0.84±0.01 | 0.85±0.02 |
| | 15 | 1.01±0.02 | 0.95±0.06 | 1.07±0.03 | 0.99±0.15 | 0.96±0.04 |
| | | 0.78±0.02 | 0.76±0.01 | 0.84±0.02 | 0.77±0.02 | 0.76±0.03 |
| | 10 | 0.85±0.03 | 0.79±0.04 | 0.79±0.03 | 0.72±0.04 | 0.82±0.08 |
| | | 0.72±0.02 | 0.65±0.05 | 0.69±0.03 | 0.64±0.02 | 0.69±0.04 |
| 70 | 20 | 0.93±0.04 | 0.99±0.05 | 1.05±0.04 | 1.05±0.03 | 1.08±0.04 |
| | | 0.66±0.02 | 0.78±0.01 | 0.80±0.00 | 0.80±0.01 | 0.83±0.01 |
| | 15 | 1.00±0.06 | 0.95±0.03 | 1.03±0.02 | 0.99±0.09 | 1.04±0.05 |
| | | 0.73±0.01 | 0.72±0.01 | 0.74±0.01 | 0.73±0.03 | 0.74±0.02 |
| | 10 | 0.89±0.03 | 0.74±0.09 | 0.75±0.04 | 0.69±0.03 | 0.67±0.06 |
| | | 0.73±0.02 | 0.60±0.03 | 0.62±0.02 | 0.56±0.02 | 0.56±0.03 |

### Additional Statistical tests

This section presents the p-values of a post-hoc analysis followed up as pairwise comparisons of different days. The main goal is to follow-up Friedman test and identify the specific pairs that result in statistically significant differences which are generally not captured by a global test. The test of this choice is Wilcoxon with Bonferroni correction. The aim to identify the bottlenecks and provide suitable benchmark for any further studies that use this design.

**Figure S3.**
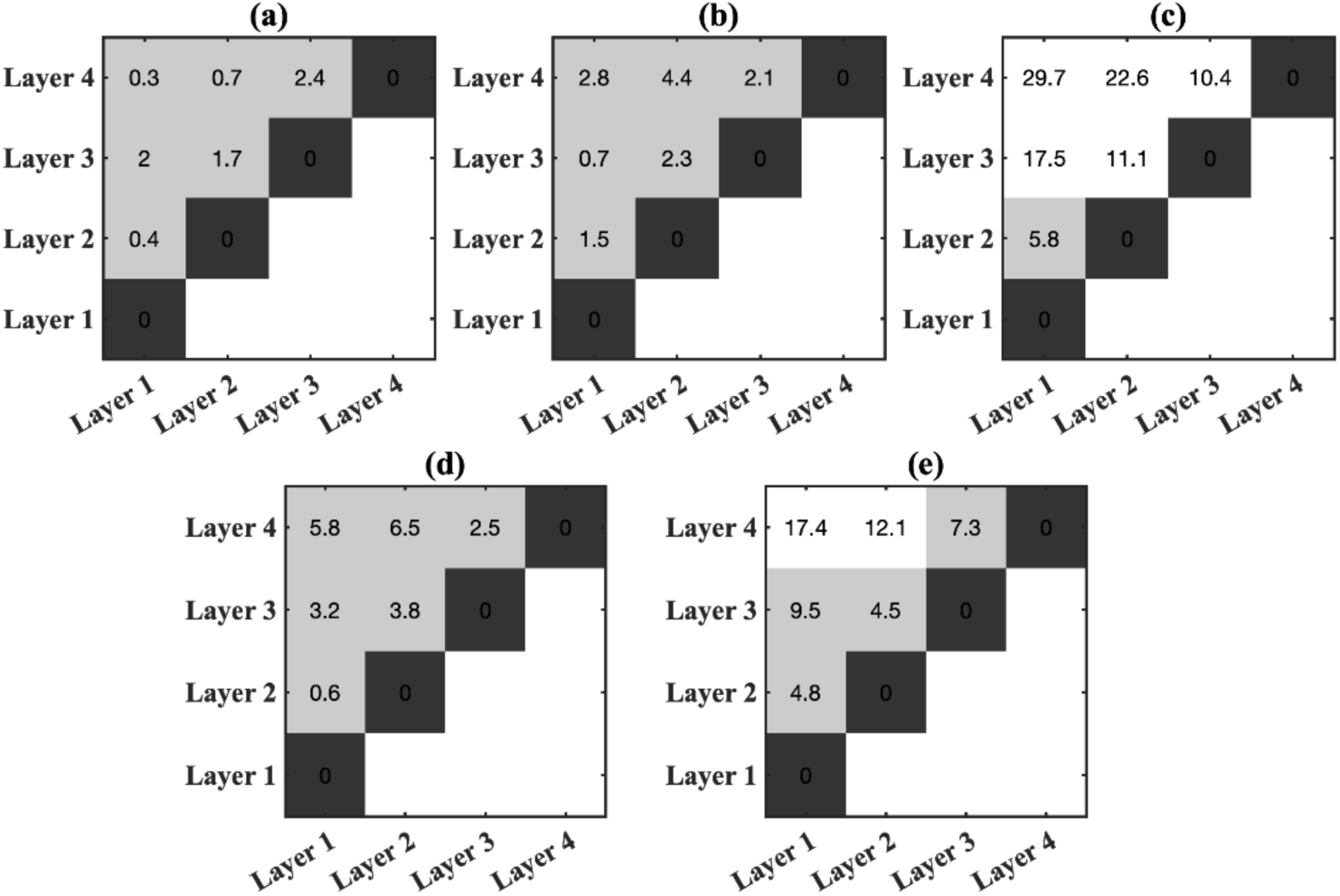
Inter Layer Pairwise *SUV_Mean_* Percentage variability matrix for (a) Day 0, (b) Day 1, (c) Day 4, (d) Day 5, (e) Day 6 – White represents the Paris that fail the 10% threshold

**Figure S4.**
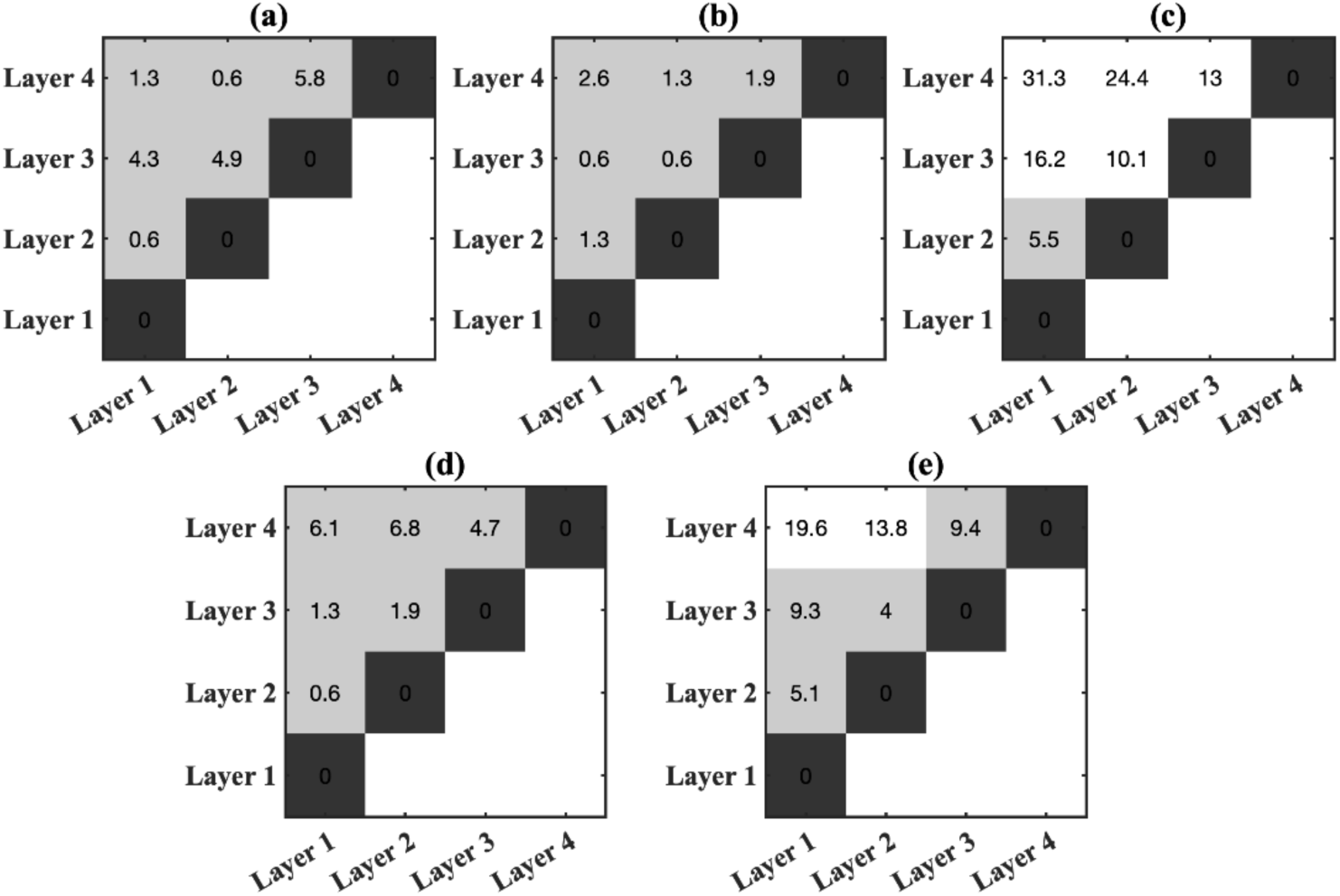
Inter Layer Pairwise *SUV_Max_* Percentage variability matrix for (a) Day 0, (b) Day 1, (c) Day 4, (d) Day 5, (e) Day 6 – White represents the pairs that fail the 10% threshold

**Figure S5.**
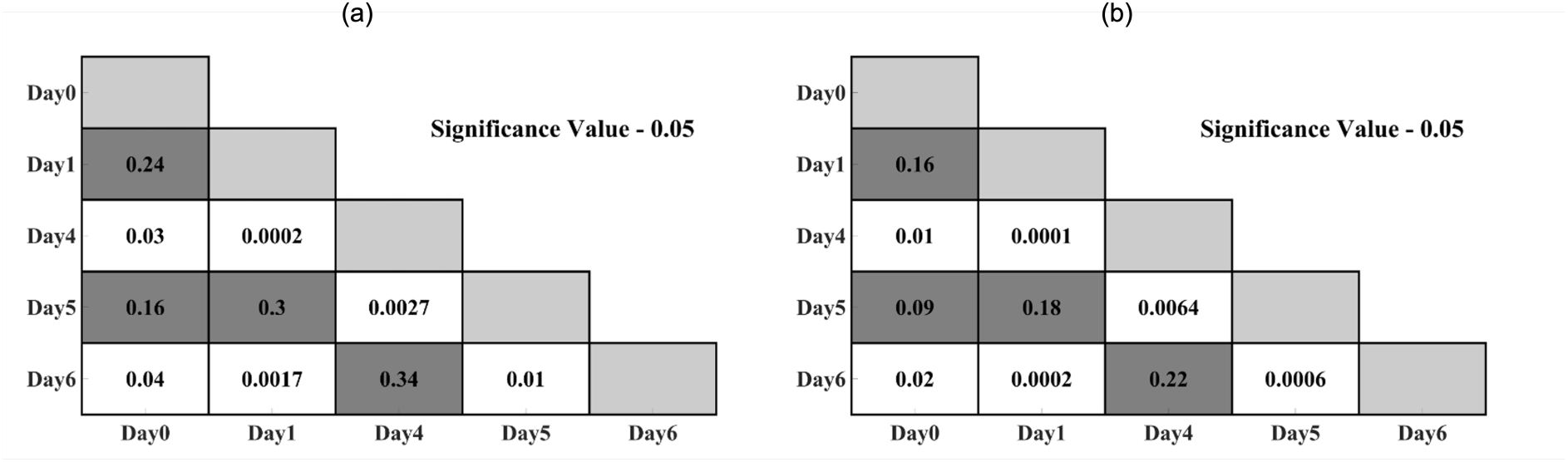
Wilcoxon signed-rank test p – value matrix for (c) TRI_max_ and (d) TRI_mean_ - White represents combinations that have statistical differences

